# COVID-19: Recovering estimates of the infected fatality rate during an ongoing pandemic through partial data

**DOI:** 10.1101/2020.04.10.20060764

**Authors:** Matteo Villa, James F. Myers, Federico Turkheimer

## Abstract

In an ongoing epidemic, the case fatality rate is not a reliable estimate of a disease’s severity. This is particularly so when a large share of asymptomatic or pauci-symptomatic patients escape testing, or when overwhelmed healthcare systems are forced to limit testing further to severe cases only. By leveraging data on COVID-19, we propose a novel way to estimate a disease’s infected fatality rate, the true lethality of the disease, in the presence of sparse and partial information. We show that this is feasible when the disease has turned into a pandemic and data comes from a large number of countries, or regions within countries, as long as testing strategies vary sufficiently. For Italy, our method estimates an IFR of 1.1% (95% CI: 0.2% – 2.1%), which is strongly in line with other methods. At the global level, our method estimates an IFR of 1.6% (95% CI: 1.1% – 2.1%). This method also allows us to show that the IFR varies according to each country’s age structure and healthcare capacity.

## Introduction

Studies on international and intra-country variation of COVID-19’s lethality abound (Sajadi et al. 2020, Bayer and Kuhn 2020, De Natale 2020, Oke and Henegan 2020, Ma et al. 2020). Indeed, the ratio of dead to confirmed cases, the case fatality rate (CFR), varies widely between countries and regions of the world. For instance, as of 7 April 2020, the UK’s CFR was 11%, while Singapore’s was 0.4%. This elicits strong scientific interest, and studies have attempted to explain why this is so. However, it is widely known that the crude CFR is an imperfect, unreliable measure for how lethal a disease actually is (Famulare 2020). This is particularly the case for diseases whose severity appears to vary widely at the individual level, giving rise to a large share or asymptomatic or pauci-symptomatic carriers. In such cases, the CFR can be used as a first approximation of the likely stress imposed by an epidemic on hospitals, although in the early stages of the epidemic it, too, needs to be corrected (Nishiura et al. 2009). However, it gives close to no information on how lethal a disease actually is.

In order to have a better picture of the overall lethality of a disease it is crucial to estimate the infected fatality ratio (IFR), which is the ratio of dead to infected cases. However, while calculating the CFR is straightforward, estimating the IFR is much harder, because it requires to find plausible evidence on the overall prevalence of the disease in a given population.

Previous research on COVID-19 has relied on “natural experiments” to recover a plausible IFR estimate. In particular, it has focussed on the case of the Diamond Princess, a cruise ship that has been quarantined off the coast of Japan, and for which the number of total infected patients is known with reasonable certainty [Russell et al. 2020]. This research estimated an IFR of 1.2% (95% CI: 0.38% – 2.78%) for the people aboard the ship, consistent with an IFR of 0.5% (95% CI: 0.2% – 1.2%) for China. Another method [Verity et al. 2020] has involved obtaining age-stratified estimates by leveraging data from China and 37 other countries, informed by PCR testing of international Wuhan residents returning on repatriation flights. For China, this method recovered an IFR of 0.66% (95% CI: 0.39% – 1.33%), which is broadly in line with Russell et al. [2020].

Given that the severity of COVID-19 increases with age, and that estimates of the IFR from the two studies above have been age-stratified, these studies can be used to calculate plausible IFRs for other countries with different age structures. Ferguson et al. [2020] estimate the United Kingdom’s IFR at around 0.9% (95% CI: 0.4% – 1.4%), while Villa [2020] estimates Italy’s IFR at 1.1% (95% CI: 0.5% – 1.8%).

In this paper, we propose a method that leverages international but partial data during a pandemic in order to recover plausible estimates of the average IFR, both within and between countries. We show that, when applied to Italian regions, this method is able to recover estimates of Italy’s IFR that are in line with estimates calculated through other methods.

## Estimating the IFR in Italy: a case study

As of April 8 2020, Italy’s CFR hovered at around an implausibly high 12.7%. This contrasts with estimates for the IFR, as described above, that settle at around 1.1% for the country. We therefore set out to explain why this is so, and to propose a novel method to estimate a country’s IFR that leverages intra-country variation in testing procedures.

We collected data from Italy’s Civil Protection agency on confirmed cases, confirmed deaths, and the number of tests carried between 24 February and 26 March 2020, in each of Italy’s 20 regions.^3^ We stopped our observation period after 32 days in order to limit potential bias: the number of tests counted by Italy’s Civil Protection may include duplicate tests per person, and the latter tend to increase over time.^4^ We built a panel dataset and calculated the crude CFR and the number of cases per test, for each region and each day.

We propose to use the number of cases per test as a proxy for each Italian region’s testing strategy. The assumption is that when a testing strategy is wide, the number of positive cases per test should be expected to drop substantially, because milder or asymptomatic carriers are much harder to find and require to sample a larger share of the population, whose test results are expected to be negative. Vice versa, when a testing strategy is restrictive, tests are reserved to more severe cases which are much easier to spot, so that each test is expected to turn out positive with higher probability.

Our hypothesis is that, since wider testing strategies end up testing a larger sample of the population, the CFR resulting from these should come nearer to approximating the IFR of a disease, while more restrictive testing strategies will lead to a higher crude CFR by missing many mild and asymptomatic cases. Figure 1 shows that this expectation appears to be confirmed when we look at variation between Italian regions.

**Figure 1.**
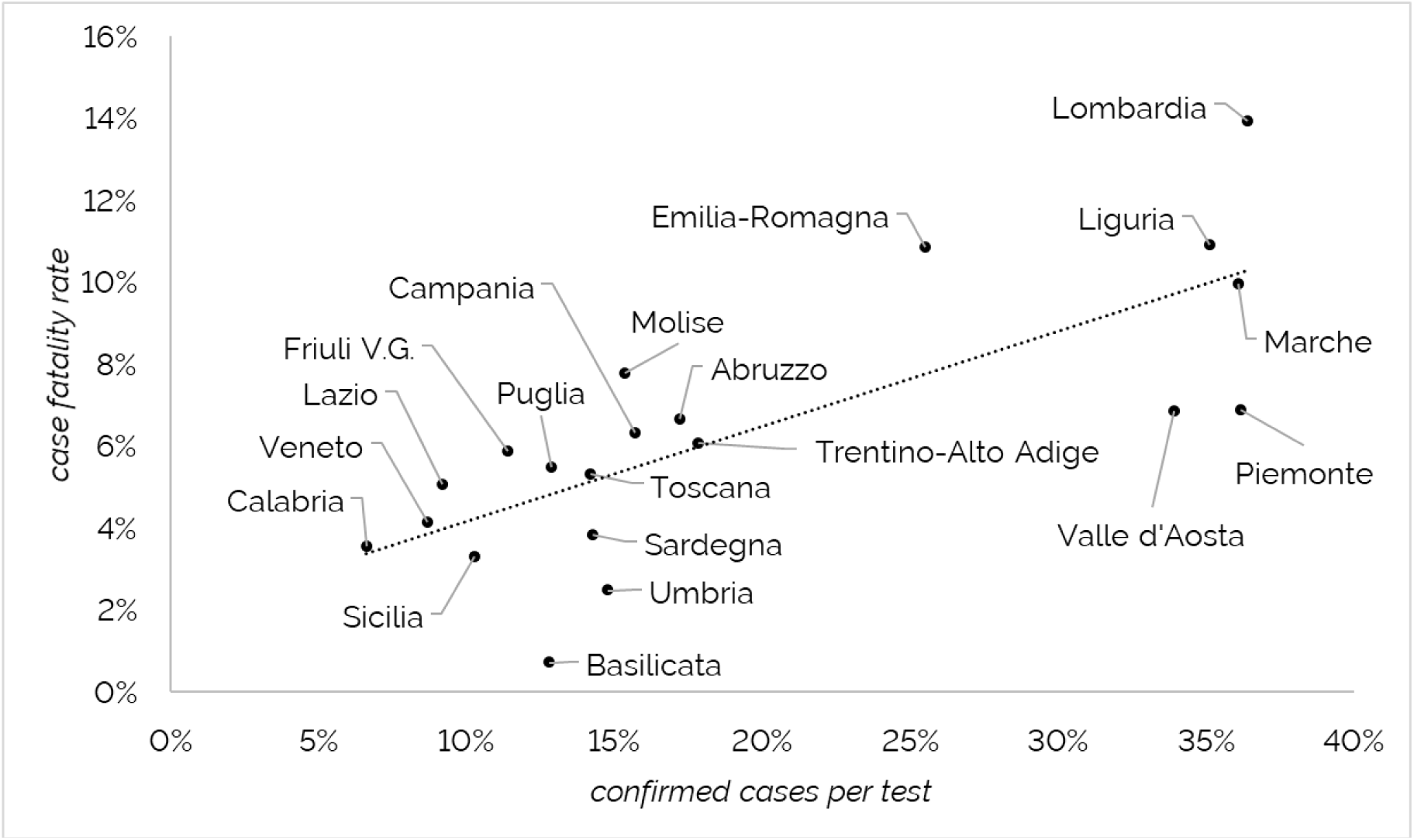
Bivariate relationship between the positive COVID-19 cases per each test carried out, and the disease’s case fatality rate, all Italian regions (as of 26 March 2020) Source: authors’ elaborations of data from the Italian Civil Protection.

We therefore propose a methodology for estimating COVID-19’s IFR by setting the number of cases per test at close to zero. This is equivalent to extrapolating COVID-19’s CFR for the widest possible testing strategy, returning a very large share of negative test results. In deriving this method, we were inspired from the Scatchard plot, a linear method used in pharmacology to estimate the affinity of a drug and the density of binding sites from partial saturation data (Scatchard 1949).

Given the characteristics of a panel data regression, we controlled for regional fixed effects, accounting for within-region variation. Moreover, on 27 February 2020 the Italian central government asked regions to comply with WHO recommendations to limit testing to symptomatic cases. This constituted a nationwide policy shock, with regions restricting their testing strategies at once – although wide regional variation remained present. Therefore, we introduced a time control for the expected increase in the CFR and cases per test as a result of this nation-wide policy change. Finally, in order to limit small-sample variation, we used the subset of region-days after cumulative deaths reached or exceeded 10 (n = 227). Given that our sample exhibits moderate heteroskedasticity, we use both a robust OLS and a robust weighted OLS in order to check whether results change significantly.

Table 1 shows our results. Models 1 and 2 show that there is a significant and substantial relationship between the testing strategy and the crude CFR, which goes in the expected direction. It also shows the importance of accounting for nationwide policy shocks and fixed effects at the regional level. Through the estimated coefficients, we recover an estimate of the Italian IFR at 1.1% (95% CI: 0.2% – 2.1%) in both models, showing that results are not affected by heteroskedasticity. While the confidence intervals for our estimates are wider than estimates calculated through other methods, our central estimate is strongly consistent with the estimate calculated in Villa [2020], which in turn was derived from Verity et al. [2020] and Russell et al. [2020].

**Table 1.** Results of robust OLS (Model 1) and robust weighted OLS (Model 2), explaining variation in regional CFRs in Italy.

|  | **Model 1** | **Model 2** |
| --- | --- | --- |
| Cases per test | 7.9 (2.6) ** | 7.6 (2.4) ** |
| Nationwide policy shock | .26 (.03) *** | .27 (.02) *** |
| (constant) | 1.15 (.50) * | 1.15 (.46) * |
| Regional fixed effects | YES | YES |
| *R squared* | 0.855 | 0.866 |
| *N* | 227 | 227 |

Robust standard errors in parentheses.
Significance levels: \* = .95 \*\* = .99 \*\*\* = .999

## Estimating the IFR at the international level

In an attempt to recover plausible estimates for COVID-19’s IFR at the global level, we deployed our method in a dataset with global data on confirmed deaths, confirmed positive cases and tests carried out. Our data is a cross section of the 58 countries that had recorded at least 20 confirmed deaths of COVID-19 patients as of 8 April 2020 (see Figure 2).

**Figure 2.**
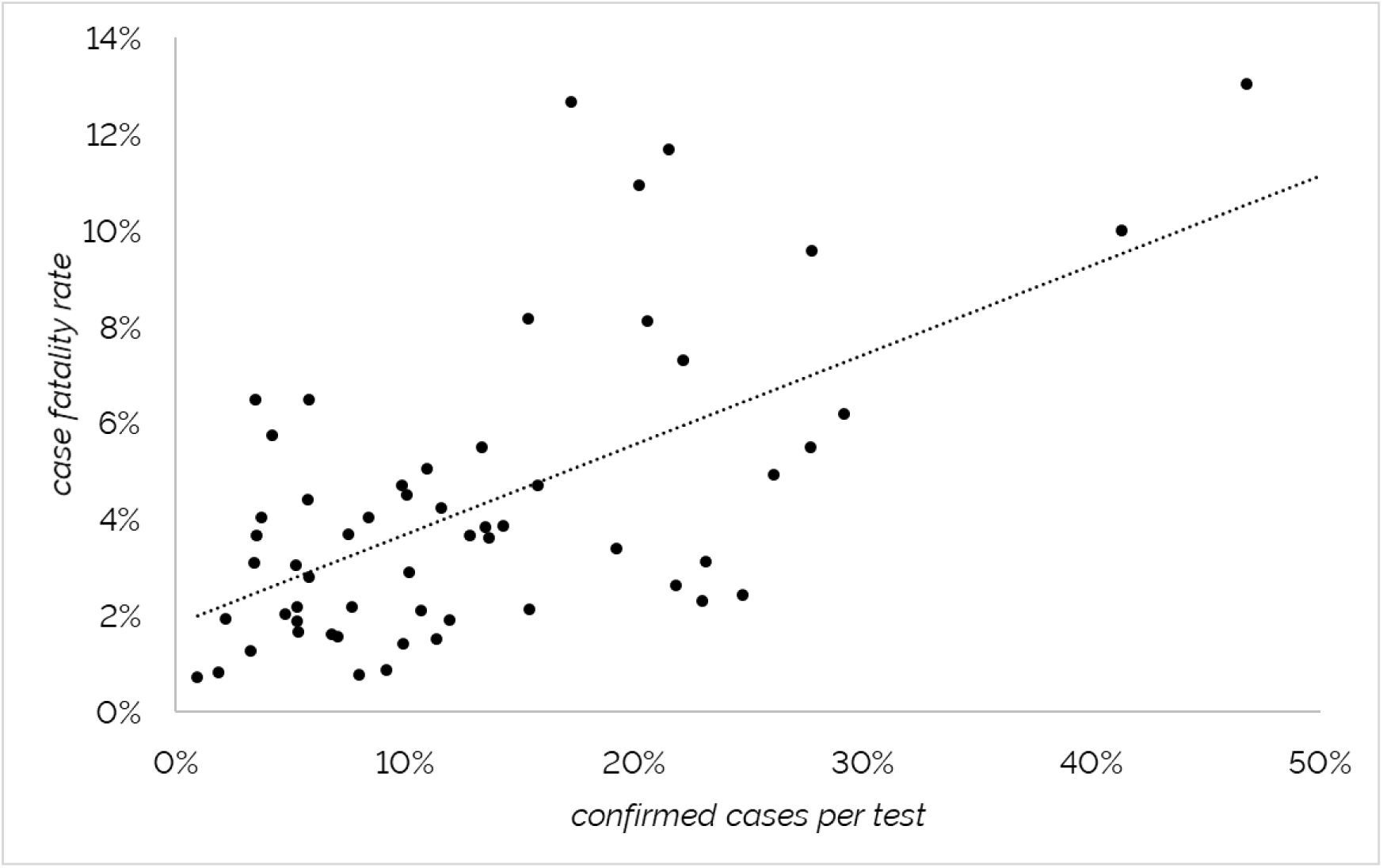
Bivariate relationship between the positive COVID-19 cases per each test carried out, and the disease’s case fatality rate, 58 countries (as of 8 April 2020) Source: authors’ elaborations of data from countries’ official authorities.

Given wide variability in age structure, healthcare systems, wellbeing, and regime type, we included the additional controls described in Table 2. Again, we estimated a weighted least squares regression with robust standard errors. Table 3 reports our results.

**Table 2.** Descriptives of control variables for the regression at the global level.

| **Control (year, source)** | **Range** | **Avg (std. dev.)** |
| --- | --- | --- |
| Population ages 65 and above<br>(2018, World Bank) | 3.3% – 27.6% | 14.2% (6.0%) |
| Current health expenditure, %<br>GDP (2016, World Bank) | 2.8% – 17.1% | 7.8% (2.7%) |
| Democracy (2019, Aggregate<br>Freedom House score) | 17 – 100 | 74.1 (23.8) |
| GDP per capita PPP, logged<br>(2018, World Bank) | 8.5 – 11.5 | 10.1 (0.7) |
| Healthcare access and quality<br>index (Institute for Health<br>Metrics and Evaluation, 2015) | 43.1 – 91.8 | 76.2 (12.5) |

**Table 3.**
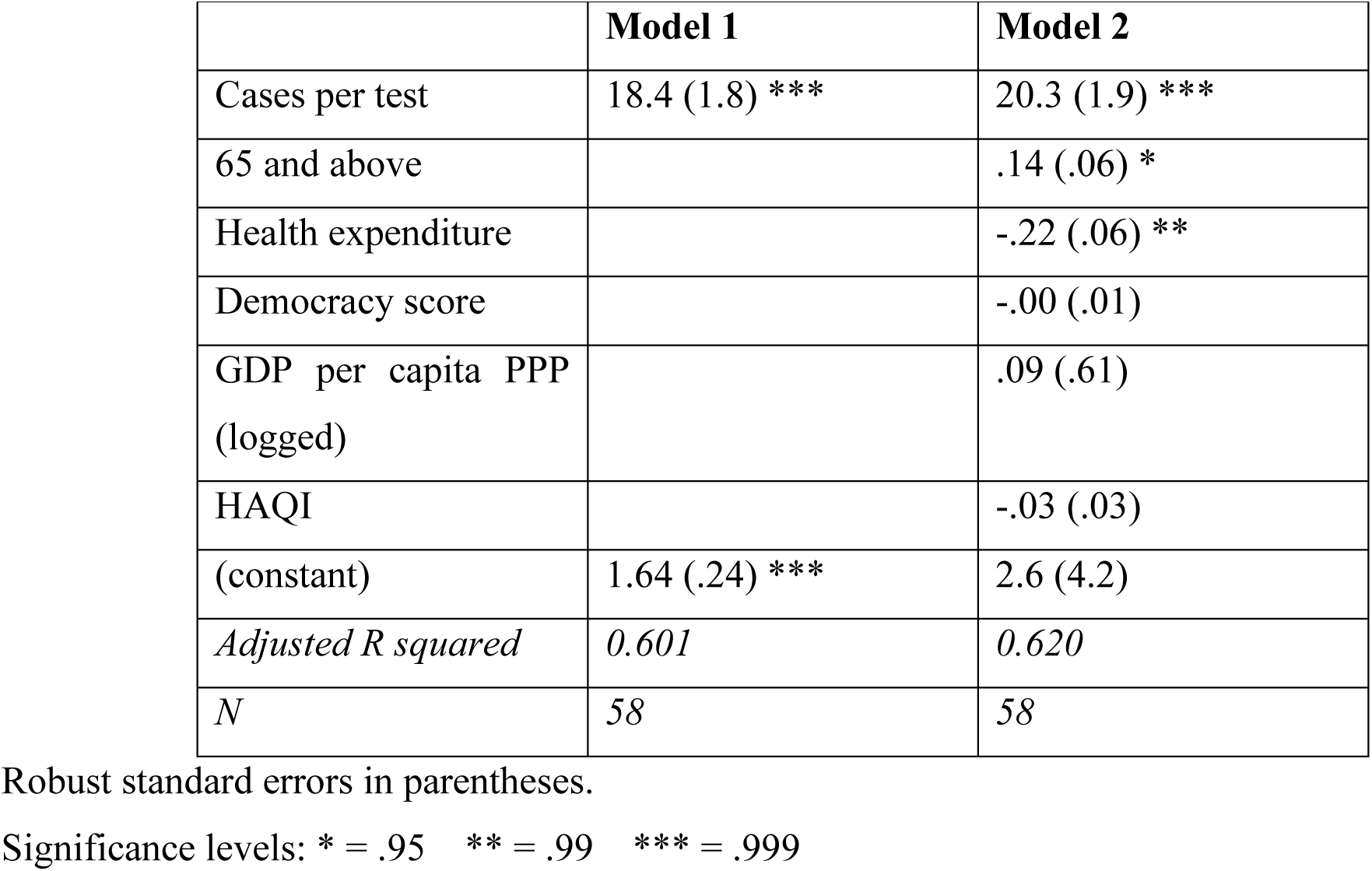
Results of robust weighted OLS, explaining variation in global CFRs.

|  | **Model 1** | **Model 2** |
| --- | --- | --- |
| Cases per test | 18.4 (1.8) *** | 20.3 (1.9) *** |
| 65 and above |  | .14 (.06) * |
| Health expenditure |  | -.22 (.06) ** |
| Democracy score |  | -.00 (.01) |
| GDP per capita PPP<br>(logged) |  | .09 (.61) |
| HAQI |  | -.03 (.03) |
| (constant) | 1.64 (.24) *** | 2.6 (4.2) |
| *Adjusted R squared* | *0.601* | *0.620* |
| *N* | 58 | 58 |

Robust standard errors in parentheses.
Significance levels: \* = .95 \*\* = .99 \*\*\* = .999

Models show that the crude CFR at the global level is strongly biased and that it strictly depends on testing capacity and strategies. In Model 2, two other variables appear to explain differences in the crude CFR: a higher proportion of persons aged 65+ significantly increases the observed CFR, while higher health expenditure significantly decreases the CFR. All results are in line with expectations, for a disease that is known to be much more lethal at older ages, and with health expenditure proxying for the preparedness of a healthcare system after controlling for age and testing strategies.

Having estimated the regression’s coefficients, we used our strategy to recover COVID-19’s IFR at the global level. As of 8 April 2020, Model 1 recovers a global IFR of 1.6% (95% CI: 1.1% – 2.1%), while after correcting for age and health expenditure Model 2 recovers a global IFR of 1.4% (95% CI: 0.8% – 1.9%). This contrasts with a crude global CFR of 6.0% as of the same date, and with a WHO estimate of 3.8% as of 28 February 2020 [WHO 2020].

Finally, we are able to model how the IFR changes as old age or health expenditures vary. Figure 3 shows that, when cases per test approach a very low value (i.e. when the CFR approximates the IFR), an increase in a country’s share of 65+ aged inhabitants brings about a substantial expected increase in the IFR, with a 5% increase in old age inhabitants increasing the IFR by 0.4%. Meanwhile, variations in health expenditure have a less steep but still substantial effect, with a 1% increase in health expenditure being associated with a 0.2% decrease in the IFR.

**Figure 3.**
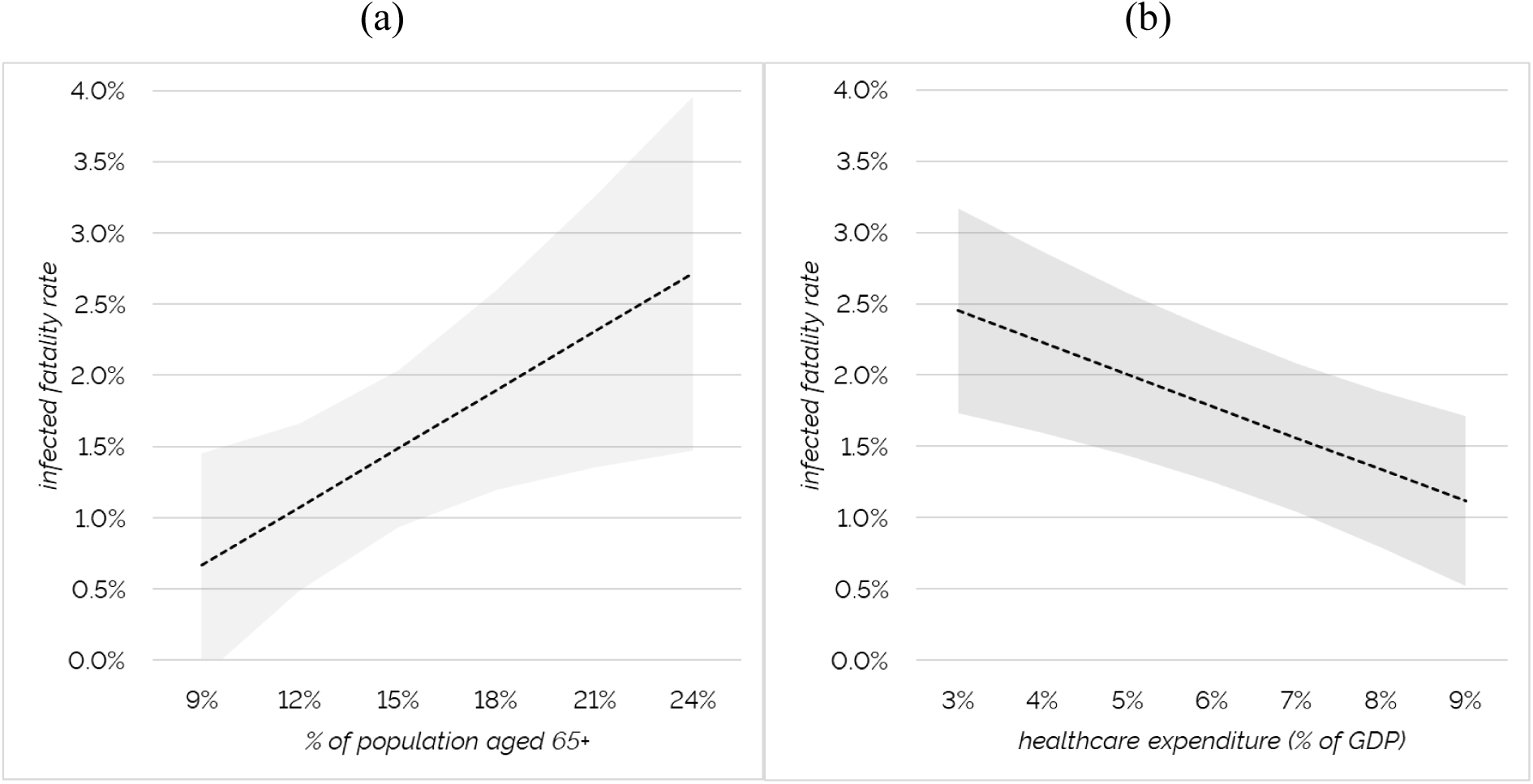
Estimated marginal effect on the IFR of (a) changes in the proportion of the population aged 65+, and (b) healthcare expenditure as a share of GDP

## Conclusion

We have shown that crude estimates of the case fatality rate are highly unreliable, and we proposed a novel method to estimate the IFR both within countries, and at the international level. We showed that variation in the CFR is strongly associated with variation in testing policies, and we estimated Italy’s and the global IFR by leveraging this knowledge.

For Italy, we recover an estimated IFR of 1.1% (95% CI: 0.2% – 2.1%) that is consistent with estimates arrived at by previous research through different methods. At the global level, we recover a global IFR of 1.4% (95% CI: 0.8% – 1.9%) and are able to explain some of its variation. Namely, the IFR is higher in countries where the proportion of inhabitants aged 65+ is larger, and it is lower in countries where health expenditure is higher.

We suggest that observers, policy makers, and international institutions take up this methodology in order to track more precisely the evolution of the current COVID-19 global pandemic and future ones.

## Data Availability

Data to replicate this study will be provided upon publication.

## Footnotes

3 In our dataset the number of regions is 21, because the autonomous provinces of Bolzano and Trento are counted separately rather than as the single region of Trentino-Alto Adige.

4 Namely, it includes tests carried out for different purposes, both to ascertain whether patients are infected and whether they have recovered. For regions providing both the number of tests carried out and the number of persons tested, we found that the difference between the two measures rises from just 1% at the start of our sample to close to 10% at the end of our observation window.

